# A Tutorial on Automated Classification of Eye Diseases Using Deep Learning

**DOI:** 10.64898/2026.03.02.26347443

**Authors:** Leila Benarous

## Abstract

Sight is one of the five senses essential to human experience, and the eyes are vital organs that require careful protection. These organs are also susceptible to a variety of diseases, some of which may develop without obvious external symptoms, necessitating specialized imaging and diagnostic techniques. Conversely, other conditions present visible signs that can be observed directly. This paper presents a practical approach to the identification of thirteen well-known eye diseases-cataract, corneal neovascularization, corneal ulcer, dry eye, endophthalmitis, globe rupture, Graves’ ophthalmopathy, ptosis, scleritis, strabismus, stye, uveitis, and xanthelasma-based on visual symptoms. Using transfer learning with the ResNet152V2 deep learning model, we demonstrate an average validation accuracy of 98.8%. The methodology is presented in a reproducible, step-by-step format suitable for educational purposes, allowing opticians, general practitioners, and learners to explore automated eye disease diagnosis. All code, datasets, and procedures are documented to facilitate practical learning and replication.

## 1. Introduction

Eye diseases encompass a broad spectrum, with complications that can range from mild inconveniences to severe conditions that threaten vision. The diverse nature of these diseases means that their symptoms can vary significantly, complicating the diagnostic process. Ophthalmologists traditionally rely on both visual symptoms and advanced imaging technologies to diagnose eye conditions accurately. Despite these tools, the complexity of eye diseases can sometimes lead to diagnostic challenges. Similarities between symptoms of different conditions, coupled with the potential for hesitation or a lack of experience among medical practitioners, can result in delayed or incorrect diagnoses. Such misdiagnoses can lead to inappropriate treatments, which not only fail to address the underlying issue but can also pose additional risks to the patient’s health.

In light of these challenges, there has been increasing interest in integrating computer vision technologies into medical diagnostics, with a particular focus on ophthalmology. Researchers are exploring how these technologies can enhance diagnostic accuracy and efficiency. By leveraging deep learning algorithms, it is possible to improve the detection and classification of eye diseases based on visual symptoms, which can be particularly beneficial for identifying early signs that might otherwise go unnoticed.

In this paper, we present a deep learning approach to diagnose fourteen specific eye diseases from visual symptoms. Our objective is to enable prompt and accurate detection of these conditions, including their early signs, thereby helping to preserve patients’ vision and improve overall treatment outcomes. This work represents a significant step towards augmenting traditional diagnostic methods with advanced computational tools.

The structure of this paper is as follows: Section 2 provides an overview of the current state-of-the-art in applying computer vision to eye disease diagnosis, highlighting recent advancements and methodologies. Section 3 details the development phases of our contribution, starting with an explanation of the deep learning model employed, specifically the ResNet152V2 architecture. We then describe the dataset preparation process and outline the training and evaluation phases of our model. Finally, Section 4 summarizes the findings and conclusions of our study, discussing the implications and potential future directions for research in this field.

## 2. Related work

Multiple researchers have leveraged computer vision for medical diagnosis due to its accuracy and rapid inference capabilities. For instance, the authors of [1] utilized convolutional neural network (CNN)-based models, specifically VGG16 and InceptionV3, pretrained on the ImageNet dataset to address multi-class diabetic eye diseases. Their study achieved a maximum accuracy of 88.3% for multi-class classification and 85.95% for mild multi-class classification using VGG16. They relied on datasets annotated by ophthalmologists to train their models. Similarly, the authors of [2] employed fundus images to train their models for diagnosing three specific eye diseases: diabetic retinopathy, glaucoma, and age-related macular degeneration. They used residual networks—ResNet50, ResNet101, and ResNet152—and found that data augmentation combined with the ResNet50 model yielded the best results, achieving an accuracy of 76.71% in classifying healthy versus diseased eyes.

Another study by authors of [3] trained their CNN model on optical coherence tomography (OCT) images to detect three eye diseases: choroidal neovascularization, diabetic macular edema, and drusen. They reported a high validation accuracy of 83.66%. Authors of [4] utilized retinal images from the STARE dataset, encompassing 14 diseases and a normal retina, leading to a total of fifteen classes. Their study involved resizing images to resolutions of 61 × 71, 46 × 53, and 31 × 35 pixels. The CNN model demonstrated the highest training accuracy at resolutions of 61 × 71 and 31 × 35 pixels, with testing accuracy peaking at 80.93% for the 31 × 35 pixel images.

In a different approach, the author of [5] implemented a non-supervised CNN model combined with a deep-belief network and SoftMax layer classifier to detect glaucoma. Using a dataset of 1200 retinal fundus images—600 from normal eyes and 600 from glaucoma patients—the model achieved an impressive accuracy of 99%. The study by authors of [6] focused on diagnosing diabetic retinopathy using a care approach that integrated the VGG19 image classification model with a mathematical model. This approach, which used CNN to enhance the dataset with additional trainable parameters, achieved a high testing accuracy of 88%.

Authors of [7] investigated cataract detection using CNN and Support Vector Machine (SVM) models. They compared these models using images of healthy eyes and cataract patients, finding that SVM outperformed CNN with an accuracy of 87.5% compared to CNN’s 85.42%. Meanwhile, authors of [8] proposed a deep learning approach to detect four types of diabetic eye diseases—cataract, diabetic retinopathy, diabetic macular edema, and glaucoma—using a dataset of 1228 fundus images. They tested five deep learning models: EfficientNetB0, VGG16, ResNet152V2, ResNet152V2 + Gated Recurrent Unit (GRU), and ResNet152V2 + Bidirectional GRU (Bi-GRU). Their results indicated that EfficientNetB0 was the most effective, achieving an accuracy of 98.76%.

Authors of [9] explored deep learning techniques for evaluating dry eye severity using corneal fluorescein staining images from two hospitals. Additionally, authors of [10] reviewed various deep learning techniques, including CNNs, Transfer Learning, Generative Adversarial Networks (GANs), Recurrent Neural Networks (RNNs), attention mechanisms, and explainable deep learning, for eye disease diagnosis. The review underscored the importance of collaboration between healthcare professionals and deep learning researchers to enhance the accuracy of eye disease diagnosis.

Building upon these advancements, our study employs deep learning residual networks to diagnose thirteen eye diseases based on visual symptoms rather than fundus images. Our aim is to aid doctors in diagnosing these diseases from early visual signs, enhancing the timeliness and accuracy of diagnoses. To position our work within the context of existing research, we have summarized the relevant state-of-the-art studies and our proposed contribution in Table 1. The following section will detail the training and evaluation processes of our approach.

**Table 1.** State-of-art solutions recap.

| Work | Dataset nature | Used model | Accuracy | Classes |  |
| --- | --- | --- | --- | --- | --- |
|  |  |  |  | Number | names |
| [1] | Fundus images | VGG16<br>InceptionV3 | VGG16: 88.3% and<br>85.95% per class | 2 | multi-class diabetic eye<br>disease and mild multi-class diabetic eye disease |
| [2] | Fundus images | ResNET 50,<br>ResNET101<br>ResNET152 | ResNET50: average<br>accuracy: 76.71% | 4 | Healthy eyes, diabetic<br>retinopathy, glaucoma, and<br>age-related macular<br>degeneration |
| [3] | OCT images | CNN | 83.66% | 3 | choroid neovascularisation,<br>diabetic macular edema<br>and drusen |
| [4] | Retinal images | CNN | 80.93%. | 15 | normal retina, hollenhorst<br>emboli, branch retinal<br>artery occlusion, cilio-<br>retinal artery occlusion,<br>branch retinal vein<br>occlusion, central retinal<br>vein occlusion, hemi-<br>central retinal vein<br>occlusion, background<br>diabetic retinopathy,<br>proliferative diabetic<br>retinopathy, arteriosclerotic<br>retinopathy, hypertensive<br>retinopathy, coats,<br>macroaneurysm, choroidal<br>neovascularization,<br>macular degeneration |
| [5] | retinal fundus<br>images | CNN combined with<br>deep-belief network<br>and SoftMax layer<br>classifier | 99% | 2 | glaucoma eye disease,<br>normal eye |
| [6] | Retinal images | Care approach | 88% | 2 | diabetic retinopathy,<br>healthy eyes |
| [7] | Retinal images | SVM and CNN | 87.5% and 85.42%<br>respectively | 2 | Cataract disease, healthy<br>eyes |
| [8] | fundus images | EfficientNetB0,<br>VGG16,<br>ResNet152V2,<br>ResNet152V2 + GRU,<br>and ResNet152V2 +<br>Bi-GRU | EfficientNetB0<br>accuracy: 98.76% | 4 | cataract, diabetic<br>retinopathy, diabetic<br>macular edema, and<br>glaucoma |
| [9] | corneal<br>fluorescein<br>staining images | Deep learning | Severity level<br>estimation | - | Dry eye |
| [10] | Eye images | CNNs, Transfer<br>Learning, (GANs),<br>(RNNs) | - | - | - |
| **Our**<br>**work** | Eye visual<br>symptoms<br>(colored<br>images) | ResNET152V2 | 98.8% | 13 | cataract, corneal<br>neovascularization, corneal<br>ulcer, dry eye,<br>endophthalmitis, globe<br>rupture, graves<br>ophthalmopathy, ptosis,<br>scleritis, strabismus, stye,<br>uveitis and xanthelasma |

## 3. Contribution

Eye diseases can present with similar early visual symptoms, such as puffiness and redness caused by inflammation, which may lead to diagnostic errors by less experienced ophthalmologists and general practitioners. Incorrect treatment resulting from misdiagnosis or delayed intervention due to hesitation can potentially result in vision loss. To address this issue, we aimed to contribute to the medical community by applying computer vision techniques for medical purposes. In this section, we describe our proposed eye disease classifier, including its functionality and preparation process. The classifier can be integrated into a mobile application, allowing doctors to perform live diagnoses or capture still images of the eye area. These images can be processed by the classifier to provide prompt and accurate diagnoses.

Our classifier is based on the ResNet152V2 deep learning model, trained on the ImageNet dataset. To train the classifier, we gathered medical images showing the symptoms of thirteen selected eye diseases. We then filtered out images with unclear symptoms and augmented the dataset to ensure the classifier had sufficient data for effective learning. Prior to training, we adjusted several hyperparameters and unfroze the model’s top layers to optimize its performance on our dataset. After training, we evaluated the model using various criteria, including accuracy, precision, recall, and the confusion matrix. Figure 1 summarizes the building process, and the following subsections detail the model architecture, dataset preparation, training process, and evaluation results.

**Figure 1.**
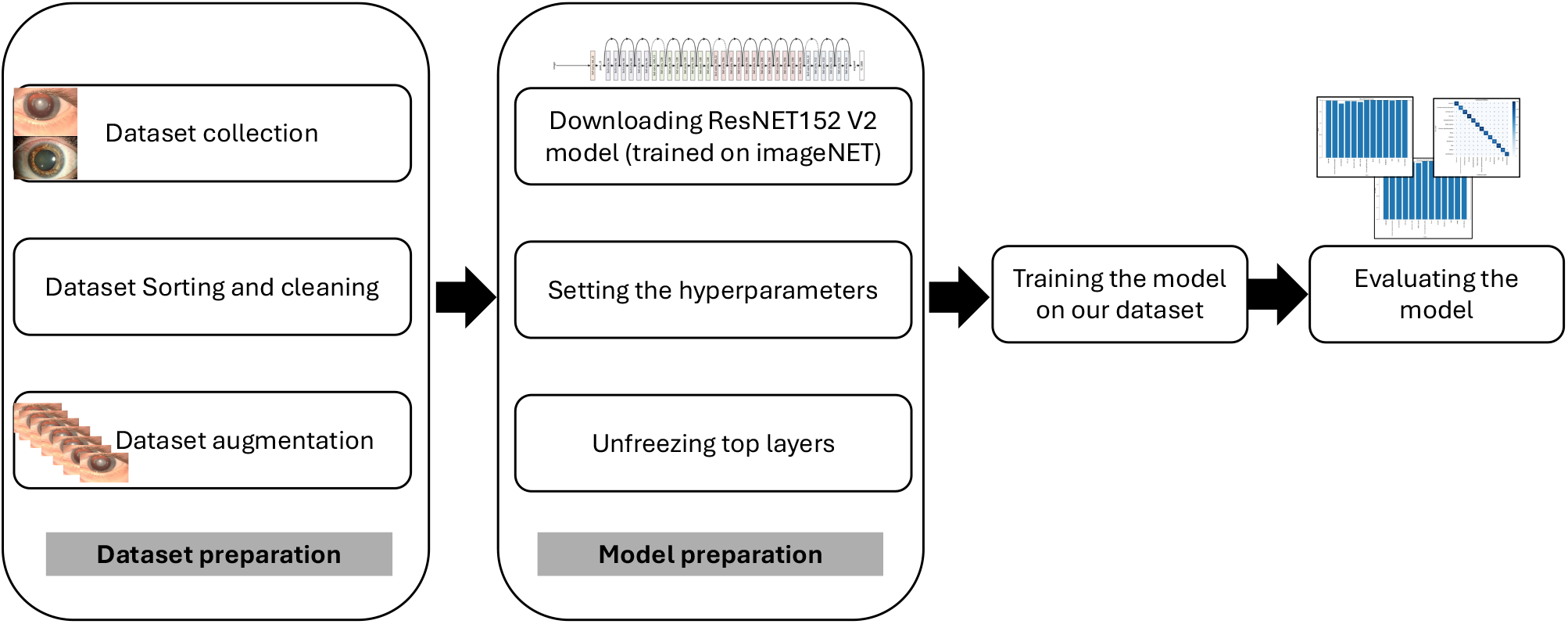
Our contribution building process

### 3.1. ResNET 152 explained

Residual networks (ResNet) are a type of convolutional neural network. The ResNet family includes several variants such as ResNet50, ResNet101, and ResNet152, with the names reflecting the number of layers in each network. In our study, we used the ResNet152V2 model, which was trained on the ImageNet dataset, containing over 14 million images across 1,000 classes. ResNet152V2 is an enhancement of ResNet152, offering improved speed and performance. Figure 2 illustrates the architecture of ResNetV2 [11]. Figure 3 shows the architecture of our model, which accepts RGB images of size 256 × 256. The model features a global average pooling layer and two fully connected layers—Dense6 and Dense7. Dense6 learns complex features, while Dense7 maps these features to class probabilities, thus handling the final classification. To prevent overfitting, a dropout layer is placed between the two dense layers.

**Figure 2.**
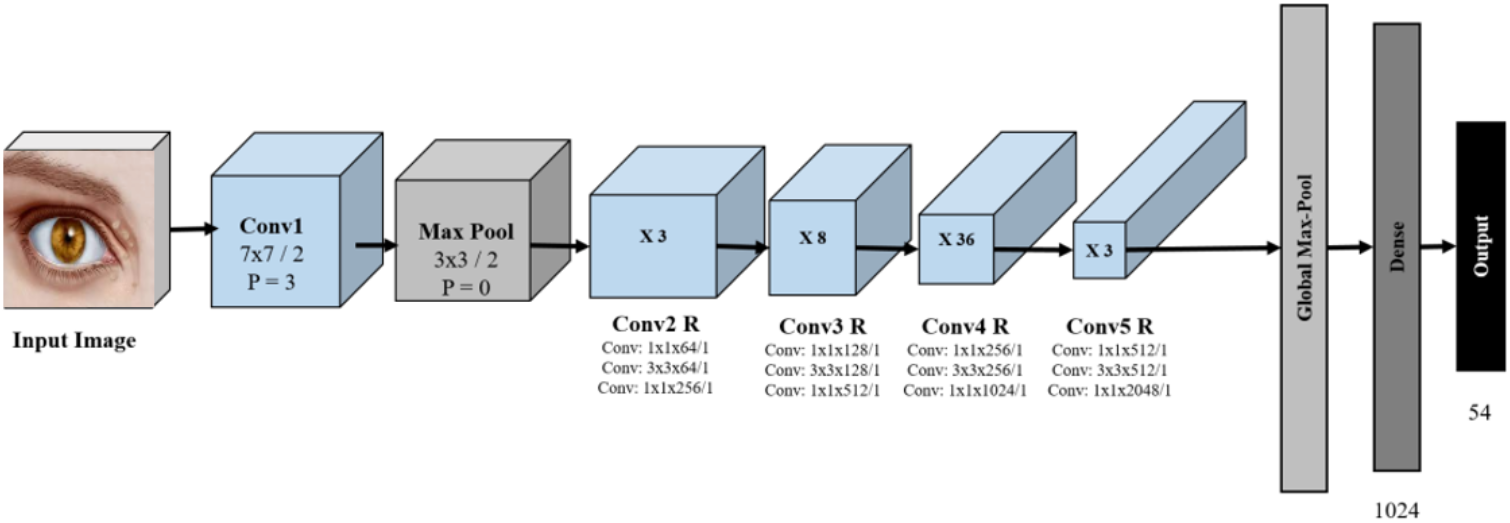
ResNET152V2 architecture *[11]*

**Figure 3.**
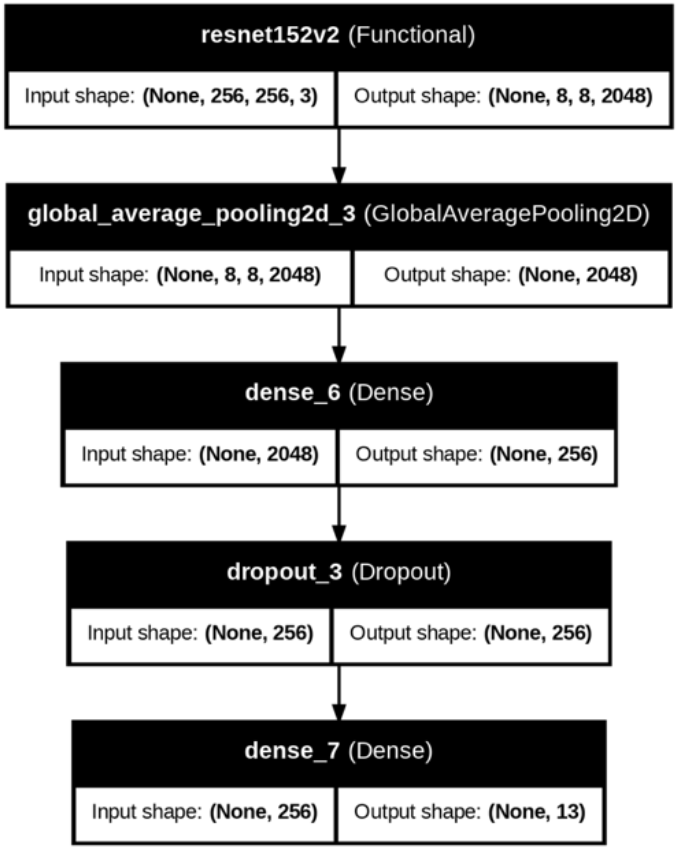
visualization of our used ResNET152V2 model

### 3.2. Dataset preparation

To enable diagnosis based on visual symptoms, we utilized Google Images to search for pictures depicting the symptoms of the thirteen selected eye diseases. Most of these images were sourced from medical websites. Table 2 summarizes the description of each disease along with its visual symptoms. After downloading and sorting the images, we discovered that the original dataset contained a limited number of pictures for each disease, which we refer to as a class. To address this issue, we needed to augment the dataset. We employed data augmentation techniques such as rotation and vertical and horizontal flipping of the images. The augmentation was performed using the Augmentor Python library. Table 3 shows the results of this augmentation, along with the original dataset for each class.

**Table 2.**
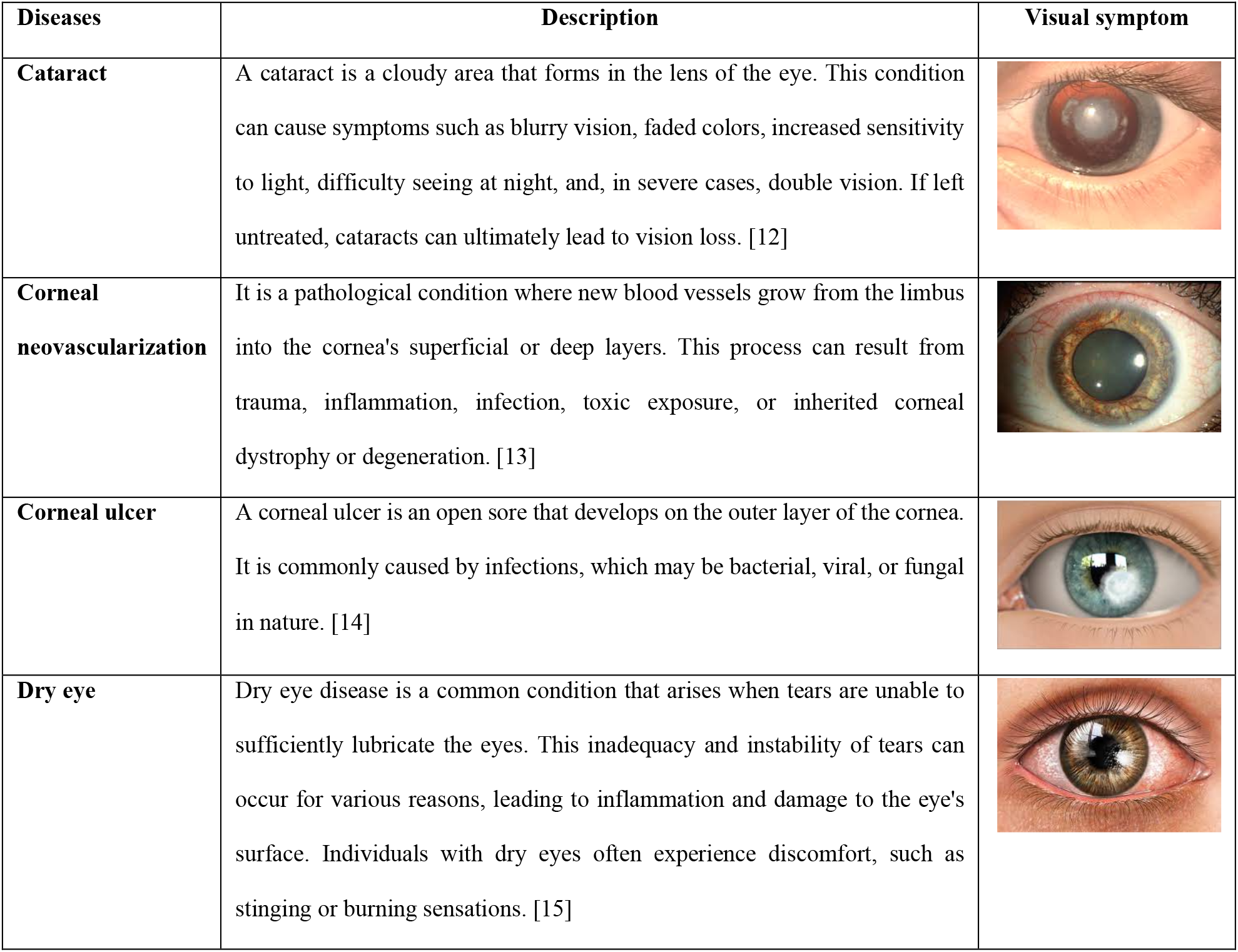
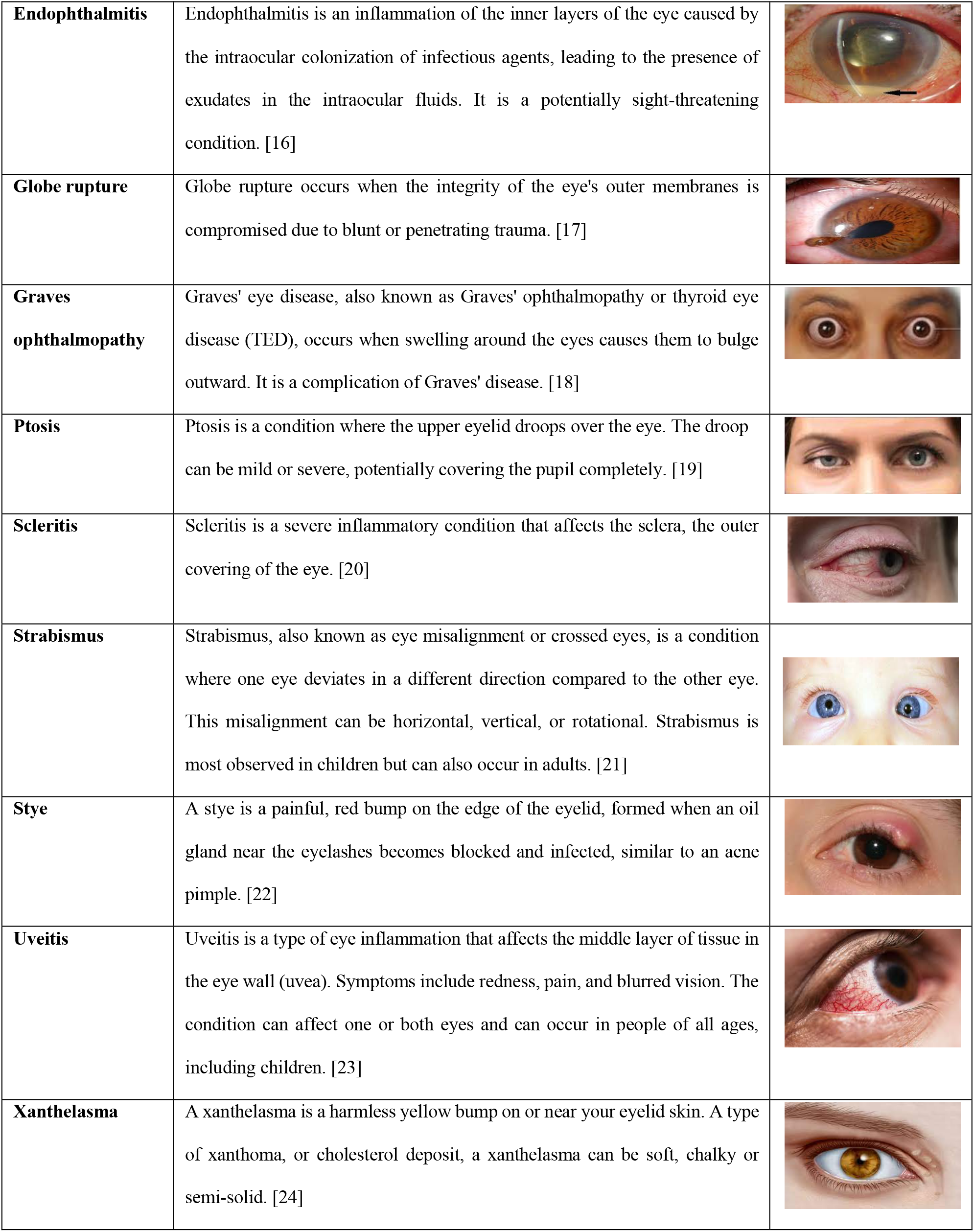
eye diseases description.

| Diseases | Description | Visual symptom |
| --- | --- | --- |
| **Cataract** | A cataract is a cloudy area that forms in the lens of the eye. This condition can cause symptoms such as blurry vision, faded colors, increased sensitivity to light, difficulty seeing at night, and, in severe cases, double vision. If left untreated, cataracts can ultimately lead to vision loss. [12] |  |
| **Corneal neovascularization** | It is a pathological condition where new blood vessels grow from the limbus into the cornea's superficial or deep layers. This process can result from trauma, inflammation, infection, toxic exposure, or inherited corneal dystrophy or degeneration. [13] |  |
| **Corneal ulcer** | A corneal ulcer is an open sore that develops on the outer layer of the cornea. It is commonly caused by infections, which may be bacterial, viral, or fungal in nature. [14] |  |
| **Dry eye** | Dry eye disease is a common condition that arises when tears are unable to sufficiently lubricate the eyes. This inadequacy and instability of tears can occur for various reasons, leading to inflammation and damage to the eye's surface. Individuals with dry eyes often experience discomfort, such as stinging or burning sensations. [15] |  |

| **Diseases** | **Description** | **Visual symptom** |
| --- | --- | --- |
| **Endophthalmitis** | Endophthalmitis is an inflammation of the inner layers of the eye caused by the intraocular colonization of infectious agents, leading to the presence of exudates in the intraocular fluids. It is a potentially sight-threatening condition. [16] |  |
| **Globe rupture** | Globe rupture occurs when the integrity of the eye's outer membranes is compromised due to blunt or penetrating trauma. [17] |  |
| **Graves ophthalmopathy** | Graves' eye disease, also known as Graves' ophthalmopathy or thyroid eye disease (TED), occurs when swelling around the eyes causes them to bulge outward. It is a complication of Graves' disease. [18] |  |
| **Ptosis** | Ptosis is a condition where the upper eyelid droops over the eye. The droop can be mild or severe, potentially covering the pupil completely. [19] |  |
| **Scleritis** | Scleritis is a severe inflammatory condition that affects the sclera, the outer covering of the eye. [20] |  |
| **Strabismus** | Strabismus, also known as eye misalignment or crossed eyes, is a condition where one eye deviates in a different direction compared to the other eye. This misalignment can be horizontal, vertical, or rotational. Strabismus is most observed in children but can also occur in adults. [21] |  |
| **Stye** | A stye is a painful, red bump on the edge of the eyelid, formed when an oil gland near the eyelashes becomes blocked and infected, similar to an acne pimple. [22] |  |
| **Uveitis** | Uveitis is a type of eye inflammation that affects the middle layer of tissue in the eye wall (uvea). Symptoms include redness, pain, and blurred vision. The condition can affect one or both eyes and can occur in people of all ages, including children. [23] |  |
| **Xanthelasma** | A xanthelasma is a harmless yellow bump on or near your eyelid skin. A type of xanthoma, or cholesterol deposit, a xanthelasma can be soft, chalky or semi-solid. [24] |  |

**Table 3.** Original and augmented dataset.

| **Diseases** | **original dataset size** | **Augmented dataset size** |
| --- | --- | --- |
| Cataract | 45 | 645 |
| Corneal neovascularization | 34 | 634 |
| Corneal ulcer | 32 | 632 |
| Dry eye | 26 | 626 |
| Endophthalmitis | 31 | 631 |
| Globe rupture | 26 | 626 |
| Graves ophthalmopathy | 32 | 632 |
| Ptosis | 23 | 623 |
| Scleritis | 20 | 620 |
| Strabismus | 42 | 642 |
| Stye | 40 | 640 |
| Uveitis | 19 | 619 |
| Xanthelasma | 35 | 635 |
| **Total** | **405** | **8205** |

### 3.3. Training and results

To diagnose the thirteen eye diseases mentioned earlier, we employed the ResNet152-V2 network weights (pretrained on ImageNet) and applied transfer learning. After downloading the model, we froze all layers except the top twenty and trained it on our augmented dataset, depicted in Section 3.2. The hyperparameters used for training are summarized in Table 4.

**Table 4.**
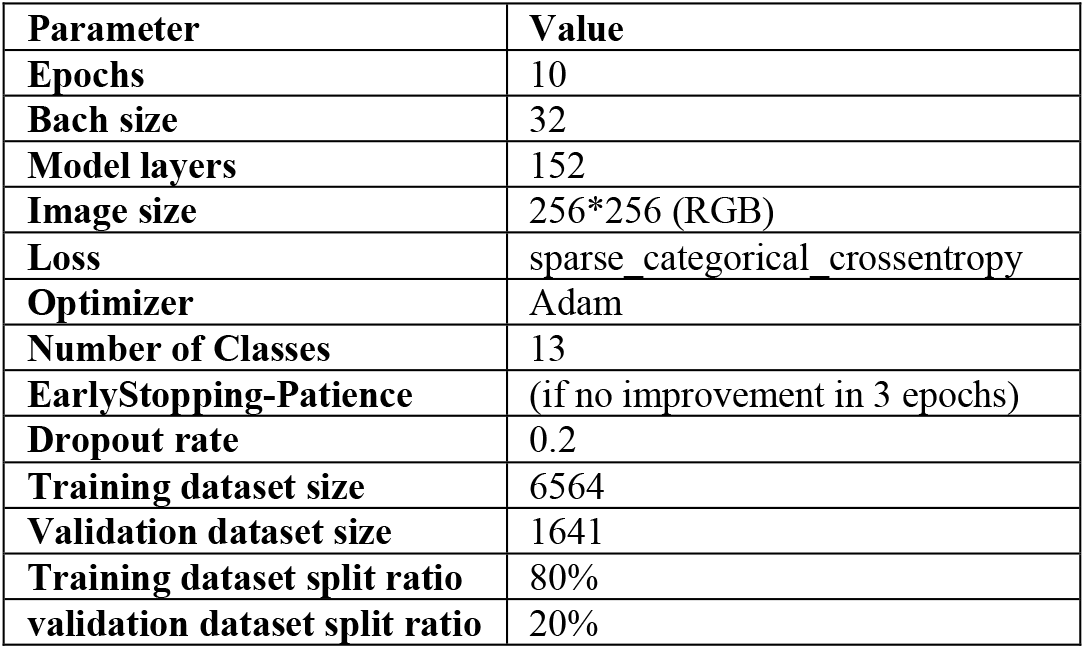
Training hyperparameters.

After training the model, we evaluated its performance. The model achieved an average classification accuracy of 98.8% for diagnosing eye diseases from their visual symptoms. Accuracy per class also remained high, ranging from 98% to 100%, as shown in Figure 4. Specifically, the model achieved 100% accuracy for six diseases: Graves’ ophthalmopathy, ptosis, scleritis, strabismus, uveitis, and xanthelasma. In addition to accuracy, we assessed the model’s performance using precision, recall, and the confusion matrix. These metrics provide a more comprehensive understanding of the model’s effectiveness, which is crucial in medical applications where minimizing false positives and false negatives is essential.

**Figure 4.**
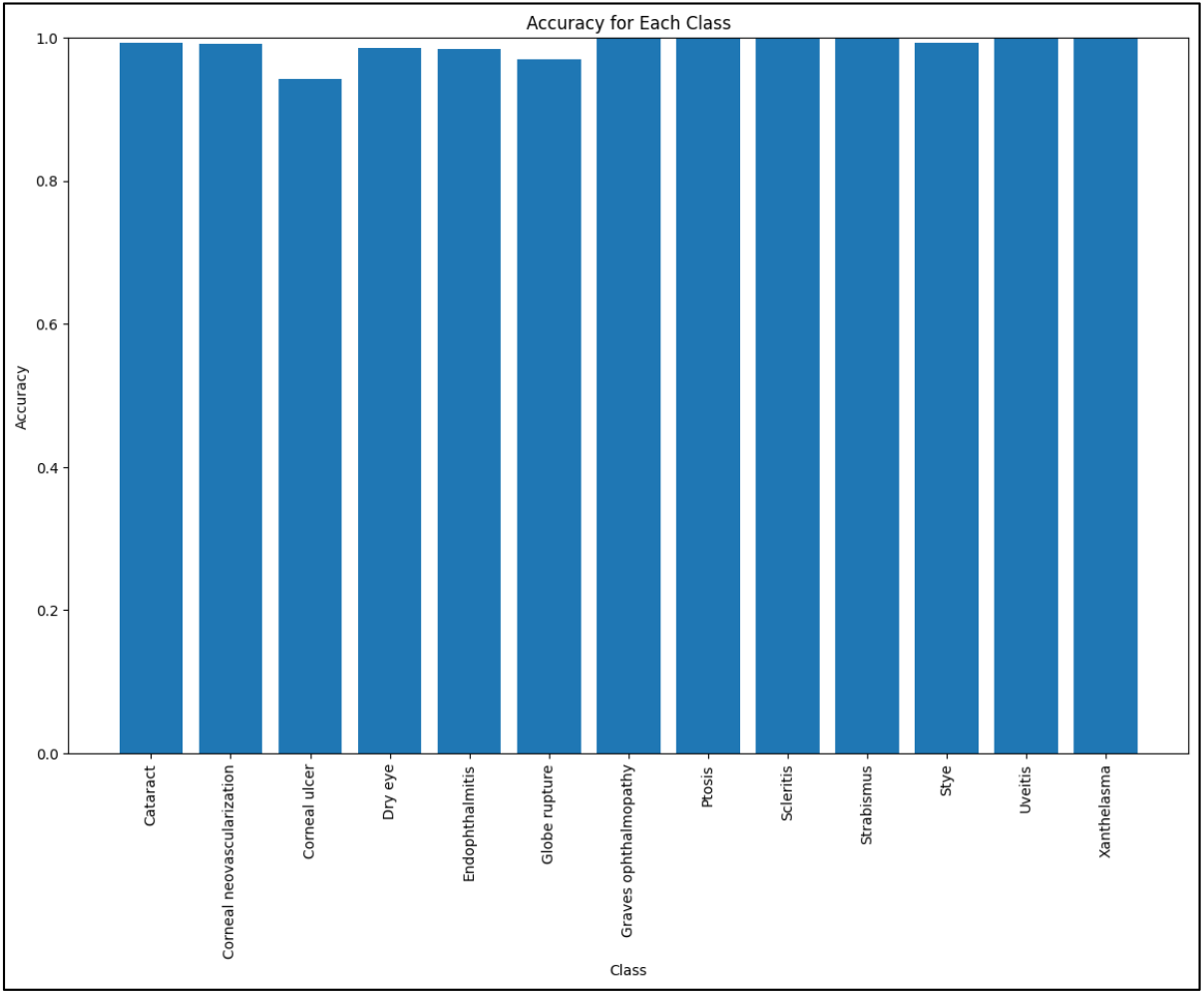
Validation accuracy per class

**Recall** is the ratio of positive samples correctly classified as positive to the total number of positive samples. It measures the model’s ability to detect actual positives. Higher recall means more positives are detected [25]. It is calculated using Equation (1). Figure 5 shows the recall ratio per class, ranging from 95% to 100%. The model achieved a 100% recall ratio for six diseases: Graves’ ophthalmopathy, ptosis, scleritis, strabismus, uveitis, and xanthelasma, indicating effective detection and accurate diagnosis of these diseases.

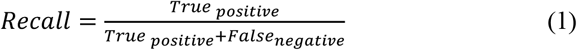

**Figure 5.**
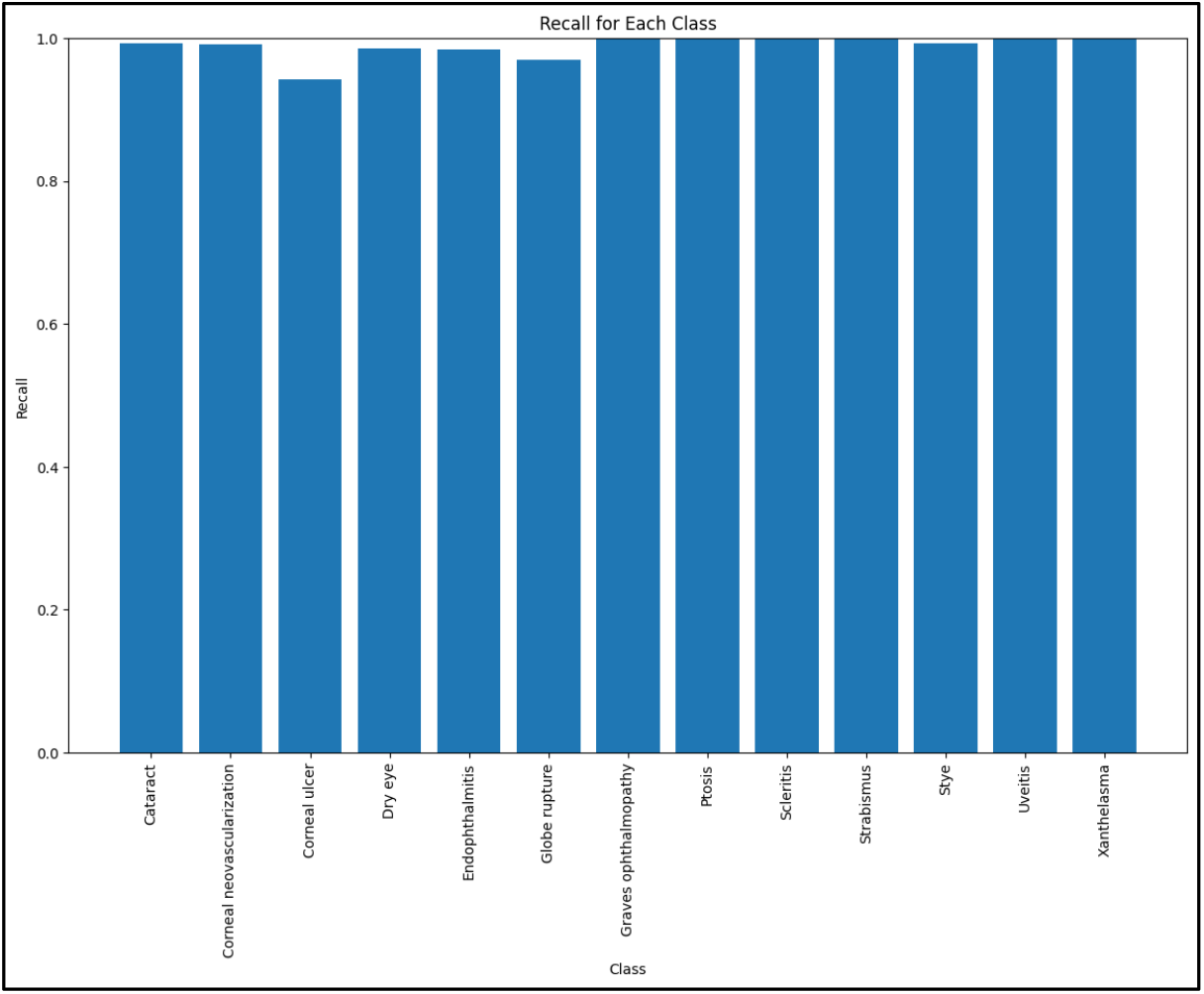
Recall per class

**Precision** is the ratio of positive samples correctly classified to the total number of samples classified as positive, whether correctly or incorrectly. It measures the model’s accuracy in identifying positive samples [25]. Precision is calculated using Equation (2). Figure 6 shows the precision for each class, ranging from 97% to 100%, with three classes—ptosis, scleritis, and strabismus—achieving a 100% precision ratio.

**Figure 6.**
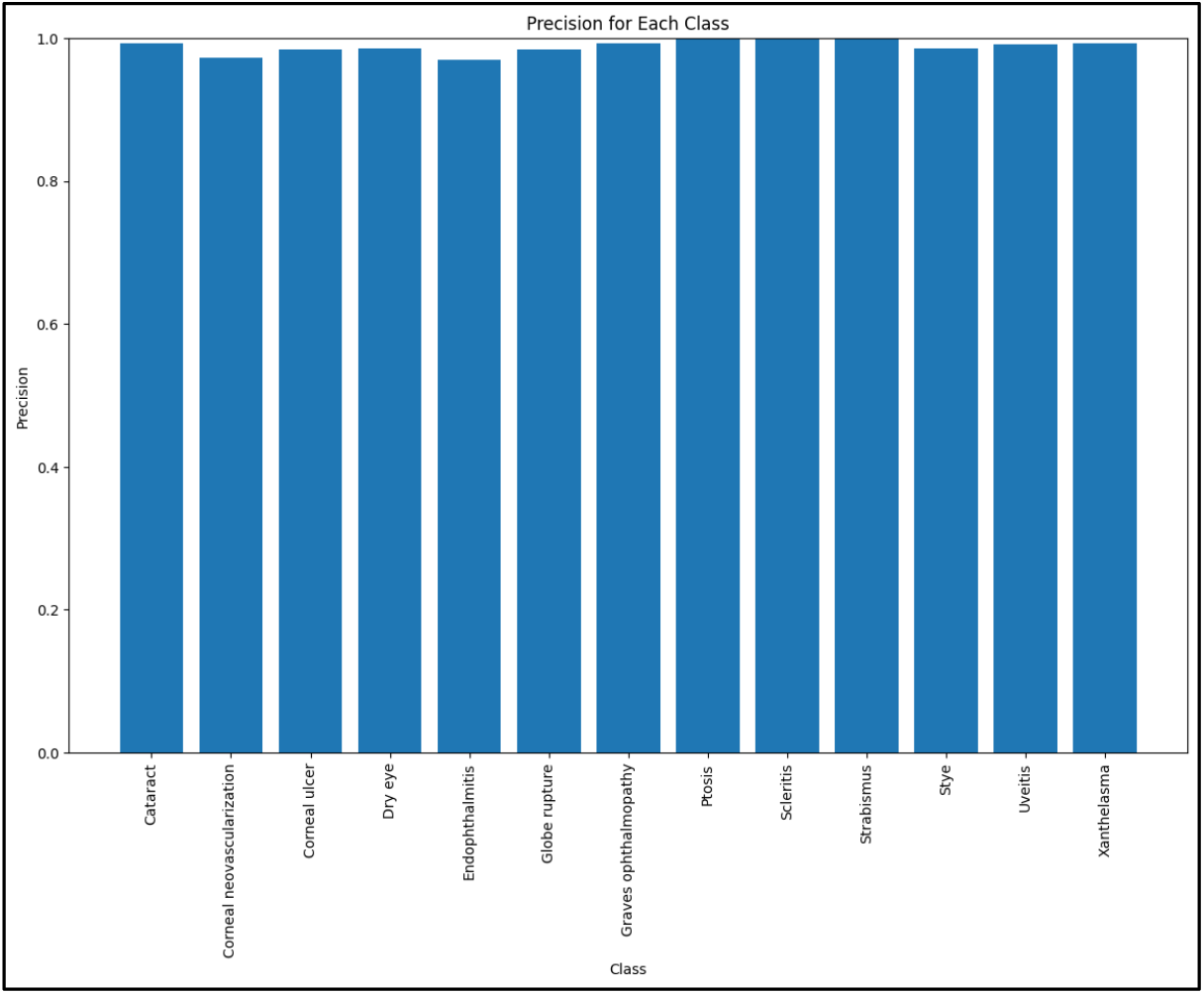
Precision per class

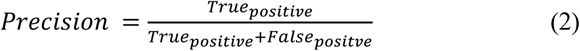

**The confusion matrix** is a tool used to evaluate the performance of a classification model by tabulating true positive, true negative, false positive, and false negative predictions. It compares actual target values with those predicted by the model [26]. Figure 7 illustrates the confusion matrix for our model, which shows overall excellent results. It demonstrates that the model classifies diseases correctly, although minor errors may occur due to similarities in visual symptoms, such as redness and puffiness, among some diseases.

**Figure 7.**
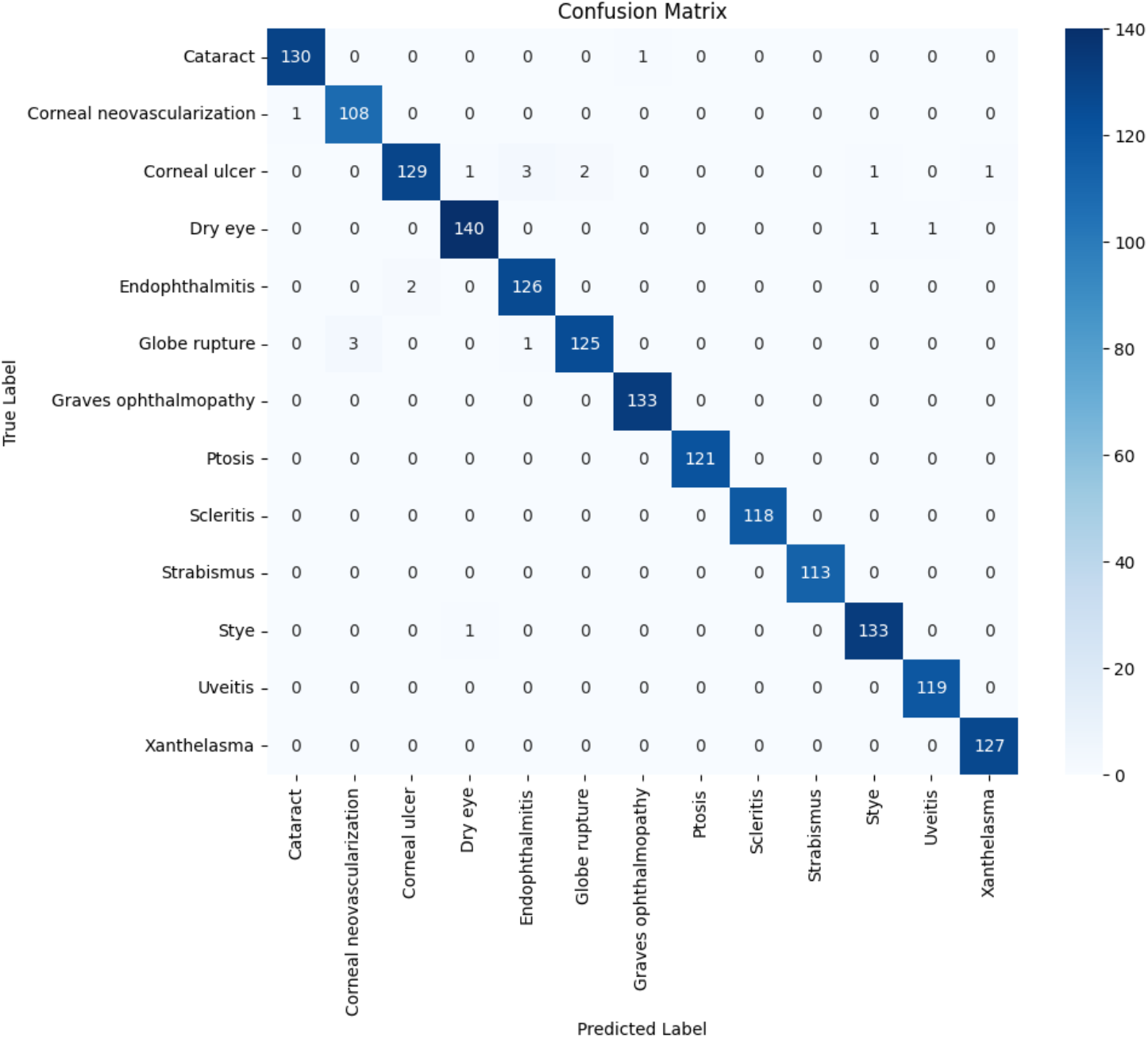
Confusion matrix

## 4. Conclusion

Eyes are a precious gift, allowing us to experience the world through vision. However, untreated eye diseases can have serious repercussions, potentially leading to vision loss. The success of treatment often hinges on accurate and timely diagnosis, which depends on identifying symptoms. Some symptoms are visible to the naked eye, while others require specialized diagnostic tools such as ophthalmoscopes, ocular ultrasound, or other imaging techniques.

To enhance both the accuracy and speed of diagnosis, many researchers have explored the use of deep learning models for disease classification. These models often rely on retinal images to detect and categorize various eye conditions. In this paper, we detail the results of training the ResNet152V2 model on color images that depict symptoms of thirteen different eye diseases. Our model demonstrated high performance, achieving accuracy, precision, and recall rates ranging from 95% to 100% for each disease class.

Looking ahead, we plan to expand our classifier to include a broader range of eye diseases. This future work will involve incorporating both retinal images and visual symptoms to improve early detection capabilities. Successful implementation of this approach will require close collaboration with ophthalmologists to obtain accurately annotated datasets, which are crucial for training robust models. Additionally, further research will be necessary to determine the most effective deep learning models, whether used individually or in combination, to handle various types of images captured by different diagnostic tools such as cameras, ophthalmoscopes, and ocular ultrasound devices. By addressing these challenges, we aim to advance the early detection of eye diseases, ultimately improving patient outcomes and preserving vision.

## Data Availability

Data are no available

## Declaration of generative AI and AI-assisted technologies in the writing process

During the preparation of this work the author used openAI chatGPT in order to proofread the paper. After using this tool/service, the author reviewed and edited the content as needed and take full responsibility for the content of the published article.

## Funding resources

This research did not receive any specific grant from funding agencies in the public, commercial, or not-for-profit sectors.

